# Fontan Subtype, Conduit Size, and Cardiac Morphologic Factors and Their Relationship to Exercise Capacity in the Fontan Circulation: A Single Ventricle Outcomes Network (SV-ONE) Study

**DOI:** 10.64898/2026.04.05.26350212

**Authors:** David M. Leone, Thomas M Glenn, Imran R. Masood, Arash A. Sabati, David A. White, Jared Hershenson, Michael Danduran, Katherine Hansen, Michael Khoury, Naomi Gauthier, Roni Jacobsen, Jesse Hansen, David Winlaw, Yves d’Udekem d’Acoz, David Morales, Alexander R. Opotowsky, the SV-ONE Investigators

## Abstract

**Background:** Exercise capacity varies among individuals with a Fontan circulation. Percent predicted peak oxygen consumption (%pVO_2_) may be influenced by ventricular morphology, Fontan subtype, and conduit characteristics, but data explaining variability in exercise capacity are limited. This study examined whether anatomical and surgical factors are associated with %pVO_2_ later in life.

**Methods:** Participants enrolled in the multicenter Single Ventricle Outcomes Network (SV-ONE) database who had cardiopulmonary exercise testing (CPET) data were included. Published reference equations were used to estimate %pVO_2_. Multivariable regression models evaluated associations between anthropometric, anatomical (diagnosis and dominant ventricle), and surgical (Fontan subtype, conduit size, and surgical era) factors and %pVO_2_. Restricted spline analyses assessed nonlinearity.

**Results:** 561 individuals with a Fontan circulation were included in the analysis; age 20 ± 8 years, 54% male, mean %pVO_2_ was 63 ± 16%. Sex and exercise modality were the strongest predictors of %pVO_2_, with females being 12% higher than males and treadmill 4.6% higher than a cycle. Age at CPET was a predictor of exercise capacity with %pVO_2_ decreasing by 0.8% per year. Ventricular morphology, diagnosis, and Fontan subtype did not have a statistical association with the primary outcome. In models restricted to patients with an extracardiac conduit (n = 330), conduit diameter and area were not associated with %pVO_2_, even after indexing to body surface area. Univariable nonlinear spline analyses suggested an optimal conduit size of 18 mm for %pVO_2_, but this was not significant after body size adjustments.

**Conclusion:** In this large multicenter cohort, surgical and anatomical features were not as important as sex, age, and body size as determinants of exercise performance in patients with a Fontan circulation. Reduced exercise capacity in this population appears to reflect progressive pathophysiological changes of the Fontan circulation rather than specific characteristics such as conduit size, ventricular morphology, or anatomy.

**CLINICAL PERSPECTIVE:** *What is new?:* In this large multicenter registry study of 561 patients with a Fontan circulation, we evaluated whether early surgical and anatomical factors, including dominant ventricular morphology, underlying diagnosis, Fontan subtype, and extracardiac conduit size, are associated with exercise performance later in life as measured by percent predicted peak VO_2_. These surgical and anatomical variables explained little of the variability in exercise capacity. In contrast, demographic and testing-related factors, particularly sex, along with age and body size, were much more strongly associated with percent predicted peak VO_2_.

*What are the clinical implications?:* Despite longstanding interest in whether Fontan configuration or dominant ventricular morphology confers superior long-term exercise capacity, this study demonstrates that these surgical and anatomical factors contribute little to exercise performance later in life. For patients with a Fontan circulation, reduced exercise capacity should not be attributed primarily to Fontan subtype or conduit size. Instead, clinical care should emphasize modifiable contributors, including regular exercise participation, preservation of skeletal muscle mass, and proactive identification and management of hemodynamic abnormalities as they evolve over time.

## BACKGROUND

With advances in surgical techniques and perioperative care, most individuals born with severe congenital cardiac conditions who undergo the Fontan procedure are now expected to reach adulthood.^1^ However, long-term functional capacity of individuals with this unique circulation depends on favorable hemodynamics.^2^ Despite advances in surgical techniques and perioperative care, long-term outcomes are affected by slow, progressive, often subclinical physiologic decline.^3,4^ Cardiopulmonary exercise testing (CPET) is used in this population to provide an empirical assessment of peak oxygen consumption (pVO_2_), functional capacity, the presence and degree of progressive subclinical physiological decline, and insights into prognosis.^4–8^

In individuals with a Fontan circulation, pVO_2_ is highly variable. Some patients achieve age-predicted values based on the general population, whereas others do not.^9,10^ Previously explored explanations for this variability include ventricular morphology, fenestration status, and other surgical factors, such as Fontan subtype.^9–17^ Conduit geometry and size have been hypothesized to influence passive pulmonary blood flow with observations suggesting that conduit size can decrease even within one year of Fontan completion.^18,19^

Prior studies have been single-center investigations around factors associated with pVO_2_ in patients with a Fontan circulation. We used multicenter registry data to examine the relationship between surgical factors from early life (conduit size and Fontan subtype), intrinsic anatomical factors (ventricular morphology and diagnosis), and individual-level factors (age at CPET, sex, exercise modality) on peak exercise capacity in adolescents and young adults with a Fontan circulation. We hypothesized that surgical factors (i.e., lateral tunnel Fontan type and conduit size) and anatomical factors (i.e., systemic right ventricular morphology) would be associated with lower percent predicted pVO_2_ (%pVO_2_).

## METHODS

### Study Design

This was a cross-sectional study utilizing pooled data from the Fontan Outcomes Network (FON), a learning health network comprised of 38 congenital heart centers in North America with the goal to improve outcomes in those living with a Fontan circulation.[REFS] As of July 2025, the Fontan Outcomes Network merged with the National Pediatric Cardiology Quality Improvement Collaborative (NPC-QIC) to form the **<u>S</u>**ingle **<u>V</u>**entricle **<u>O</u>**utcomes **<u>NE</u>**twork (SV-ONE). Data available for this study included only the most recent CPET available for each patient at the time of enrollment into the registry.

### Description of the Cohort

There were 1,114 patients enrolled within the FON between August 2022 and August 2024. The cohort included patients between the ages of 7 and 66 years who underwent CPET after their Fontan surgery (n = 663). The majority (97.1%) were performed between 2019 and 2024 (n = 545 of 561). The cohort was selected using the following inclusion criteria: (1) CPET data entered into the registry (2) Fontan surgical history/factors and anatomical factors available in the database; and (3) availability of descriptive and demographic data from the time of the CPET such as height, weight, sex, and age. Patients were excluded if the respiratory exchange ratio (RER) at pVO_2_ on the CPET was less than 1.0. An RER of 1.0 was chosen to evaluate maximal and close-to-maximal exercise tests, recognizing that many patients with a Fontan physiology often are unable to reach an RER >1.1.^15,20–22^ In addition, the **<u>F</u>**itness **<u>R</u>**egistry and the **<u>I</u>**mportance of **<u>E</u>**xercise: A National **<u>D</u>**atabase (FRIEND) cohort used for predictive values was inclusive of RER >1.0 (see below). There is also evidence that the prognostic value is preserved in individuals with acquired heart failure, even when the RER does not reach 1.1.^5,15,23,24^ Patients were also excluded if essential data, such as RER, exercise type, or pVO_2_ were missing. A schematic of cohort selection is shown in **Figure 1**.

**Figure 1.**
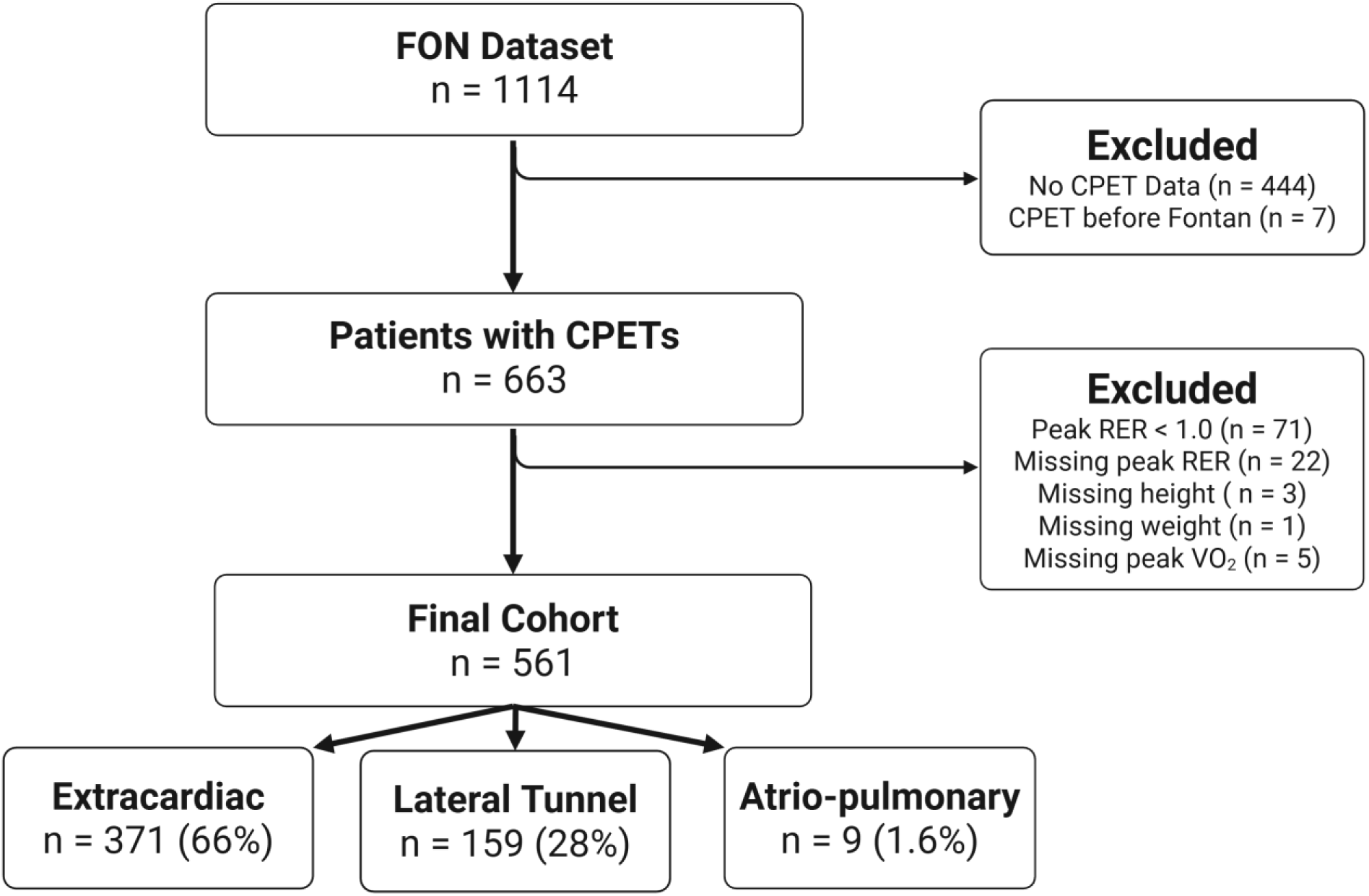
Cohort Selection. Flow diagram demonstrating derivation of the final study cohort from the Fontan Outcomes Network (FON) dataset. CPET = cardiopulmonary exercise test; RER = respiratory exchange ratio; VO_2_ = oxygen consumption.

### Data Collection and Definitions

Descriptive and demographic variables included age, sex, race/ethnicity, anthropometric data at the time of the CPET. CPET variables included test modality (cycle vs treadmill), pVO_2_ (absolute and indexed to body mass), ventilatory efficiency (VE/VCO_2_), RER at peak pVO_2_, and the ventilatory anaerobic threshold (VAT). The CPET data collection form is included in the **Supplemental Data** and is available at svone.org.

For patients ≥18 years of age at the time of the CPET, predicted pVO_2_ was determined using the age, sex, and exercise modality-specific FRIEND normative reference equation, developed from CPETs in over 10,000 healthy adult individuals.^24^

- Adults (age ≥18 years) - **FRIEND registry equation**:
  - Predicted pVO_2_ (mL/kg/min) = 45.2 − 0.35 × Age (years) − 10.9 × Sex (male = 1, female = 2) − 0.15 × Weight (pounds) + 0.68 × Height (inches) − 0.46 × Exercise Mode (treadmill = 1, cycle = 2)

For those aged <18 years at the time of the CPET, pVO_2_ was estimated using the equation from Cooper *et al*.^25^

- Youth (age <18 years) *-* Cooper et al., equations*:
  - Boys: Predicted peak VO_2_ 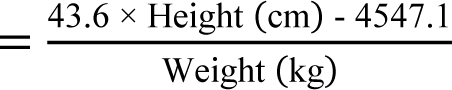
  - Girls: Predicted peak VO_2_ 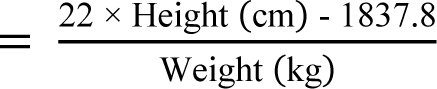

*The FRIEND equation is for predicted pVO_2_ in units mL/kg/min. To have consistent units, the Cooper equations were divided by weight in kg to calculate mL/kg/min

Percent predicted pVO_2_ was calculated as described below.

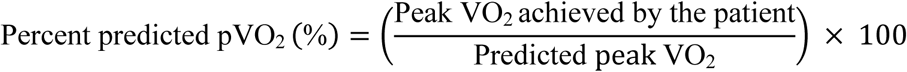

Percent predicted heart rate (%pHR) was determined using peak HR / (220 – age) × 100. The FRIEND equation was selected for adults given its established ability to risk stratify patients with a Fontan circulation and its superior prognostic performance in heart failure.^26,27^ Sensitivity analysis was conducted using the Wasserman/Hansen equation and is explained below.

### Variables of Interest

Surgical factors included Fontan subtype, Fontan surgical era, and conduit size. For Fontan subtype, patients were assigned to the following categories: 1) extracardiac conduit (ECC); 2) lateral tunnel (LT); 3) atrio-pulmonary connection (APC); or 4) other/unknown, based on categorization within the database. Conduit dimensions were described in four ways: 1) conduit size (mm); 2) indexed conduit size (mm/m^2^); 3) conduit area (mm^2^); and 4) indexed conduit area (mm^2^/m^2^). For indexed conduit measures, BSA at the time of CPET was used. When both initial and revision (*n* = 29/561; 5.2%) conduit sizes were available, the most recent value was used, provided the CPET occurred after the revision; if not, the revision data were excluded. Fontan surgical era was categorized into consecutive, approximately 8-year intervals (1980-1988, 1989-1996, 1997-2005, 2006-2013, and 2014-2022) to capture temporal trends over time. Anatomical factors included ventricular morphology where patients were assigned to the following categories: 1) right dominant; 2) left dominant; or 3) balanced/indeterminate. Due to the small sample size, patients with balanced or indeterminate ventricular morphology were grouped into a single category. Other descriptive, anthropometric, and CPET factors described above were also included for analysis.

### Statistical Analysis

Continuous variables are presented as mean ± SD for normally distributed variables, and median [25th-75th percentile] for non-normally distributed variables. The Kruskal-Wallis rank sum test was used to compare continuous variables between multiple groups with post hoc comparisons done with Bonferroni correction via the Dunn test. The primary outcome variable was %pVO_2_ achieved on CPET. Univariable and multivariable linear regression models were constructed to evaluate the relationship between predictors of interest, including demographic data, conduit size (mm), indexed conduit size (mm/m^2^), conduit area (mm^2^), indexed conduit area (mm^2^/m^2^), Fontan subtype, initial diagnosis, Fontan surgical era, and ventricular morphology. Five multivariable models were evaluated. This included the above variables, in addition to an adjustment for age, sex, and exercise modality (treadmill vs cycle ergometer). Categorical ECC groups were created to assess conduit sizes. A very small subset (*n* = 3) of those had an odd conduit size recorded, and these were rounded up to the next even integer for the categorical analysis. Semi-partial R^2^ (ΔR^2^) was computed for each individual continuous variable’s incremental impact on the model.

Restricted cubic splines were used to explore nonlinear relationships. Missing values were not imputed but were handled using pairwise deletion in regression models. The category with the largest sample size was used as the reference group for categorical variables. All analyses were conducted in R, version 4.5.0 (R Foundation, Vienna, Austria) using the RStudio environment version 2025.05.0 (Posit, Boston, MA, USA). Key packages included *rms* for regression modeling and splines, *dunn.test* for nonparametric post hoc comparisons, and *summarytools* for descriptive statistics. A two-sided p-value <0.05 was considered statistically significant.

### Ethical Considerations

This study was reviewed and considered exempt by the Cincinnati Children’s Hospital Medical Center institutional review board. Project proposal and final manuscript were approved by the FON/SV-ONE Research Working Group and Publications & Presentations committees. This study abided by the STROBE guidelines for observational studies.^28^

## RESULTS

### Study Population and Cohort Characteristics

After inclusion and exclusion criteria were applied, *n* = 561 individuals met eligibility criteria for this analysis. The mean age at time of CPET was 20 ± 8 years, with about half (*n* = 269, 48%) younger than 18 years, and 304 (54%) male. Left ventricular morphology was present in 250 (45%) patients, right in 290 (52%) patients, and balanced/indeterminate in the remainder 21 (3.7%) patients. The most common underlying diagnosis was hypoplastic left heart syndrome (*n* = 194, 35%), followed by tricuspid atresia (*n* = 101, 18%), double inlet left ventricle (*n* = 83, 15%), and unbalanced atrioventricular septal defect (*n* = 69, 12%). These results, along with less commonly occurring diagnoses can be viewed in **Table 1**.

**Table 1.** Demographics. Data displayed as n (%); mean ± SD. BSA = body surface area; AVSD = atrioventricular septal defect; DORV = double outlet right ventricle; DILV = double inlet left ventricle; HLHS = hypoplastic left heart syndrome; PAIVS = pulmonary atresia with intact ventricular septum.

| Variable (*n* = 561) | Value |
| --- | --- |
| Sex |  |
| Male | 304 (54%) |
| Age, years | 20 ± 8 |
| Age Group, years |  |
| 6-12 | 63 (11%) |
| 13-20 | 253 (45%) |
| 21-30 | 174 (31%) |
| 31-40 | 54 (9.6%) |
| ≥40 | 17 (3.0%) |
| BSA, m<sup>2</sup> | 1.67 ± 0.31 |
| Ventricular Morphology |  |
| Right | 290 (52%) |
| Left | 250 (45%) |
| Balanced/indeterminate | 21 (3.7%) |
| Fontan Type |  |
| Extracardiac Conduit | 371 (66%) |
| Lateral Tunnel | 159 (28%) |
| Atrio-pulmonary Connection | 9 (1.6%) |
| Other/Unknown | 22 (3.9%) |
| Extracardiac Conduit size, mm |  |
| ≤14 | 6 (1.7%) |
| 16 | 31 (9.0%) |
| 18 | 135 (39%) |
| 20 | 146 (42%) |
| ≥22 | 26 (7.6%) |
| Diagnosis |  |
| Balanced AVSD/DORV | 8 (1.4%) |
| DILV | 83 (15%) |
| Ebstein | 8 (1.4%) |
| HLHS | 194 (35%) |
| Mitral Atresia | 21 (3.7%) |
| Other | 52 (9.3%) |
| PAIVS | 25 (4.5%) |
| Tricuspid Atresia | 101 (18%) |
| Unbalanced AVSD | 69 (12%) |
| Fontan Era |  |
| 1980-1988 | 5 (1.0%) |
| 1989-1996 | 49 (9.5%) |
| 1997-2005 | 130 (25%) |
| 2006-2013 | 247 (48%) |
| 2014-2022 | 83 (16%) |

Most patients had an ECC (*n* = 371, 66%), followed by LT (*n* = 159, 28%), APC (*n* = 9, 1.6%), and other/unknown (*n* = 22, 3.9%). For ECC, the mean conduit diameter at time of Fontan (initial or revision) was 18.8 ± 1.7 mm. In terms of surgical era, most patients in this cohort underwent their Fontan procedure between 2006 and 2013 (*n* = 247, 48%), followed by 130 (25%) between 1997 and 2005, 83 (16%) between 2014 and 2022, 49 (9.5%) between 1989 and 1996, and 5 (1.0%) between 1980 and 1988. A summary of the demographic and clinical data are presented in **Table 1**.

### Cardiopulmonary Exercise Testing Performance

There was a near-even distribution between those tested on a cycle ergometer (273; 49%) and treadmill (288; 51%). The mean RER at peak exercise was 1.16 ± 0.10 with 146 (26%) having an RER between 1.0 and 1.1. The mean absolute pVO_2_ was 1,529 ± 490 mL/min (25 ± 7 mL/kg/min). The mean %pVO_2_ was 63 ± 16% (**Figure 2**). The VO_2_ at the VAT was 1,030 ± 357 mL/min (16.8 ± 5.4 mL/kg/min). The mean pHR was 160 ± 26 bpm (80 ± 12% predicted), and the mean oxygen saturation (SaO_2_) at peak exercise was 88.3 ± 5.7%. VE/VCO_2_ slope was 35 ± 6. Within the cohort, the mean %pVO_2_ in those 6-12 years was the highest around 70.4 ± 16.2% and was lower in those older than 21 years with a similar mean across the older 3 groups of 57-58 ± 12-20% (p = 0.011 across age groups; **Supplemental Material**). The CPET findings are summarized in **Table 2**.

**Figure 2.**
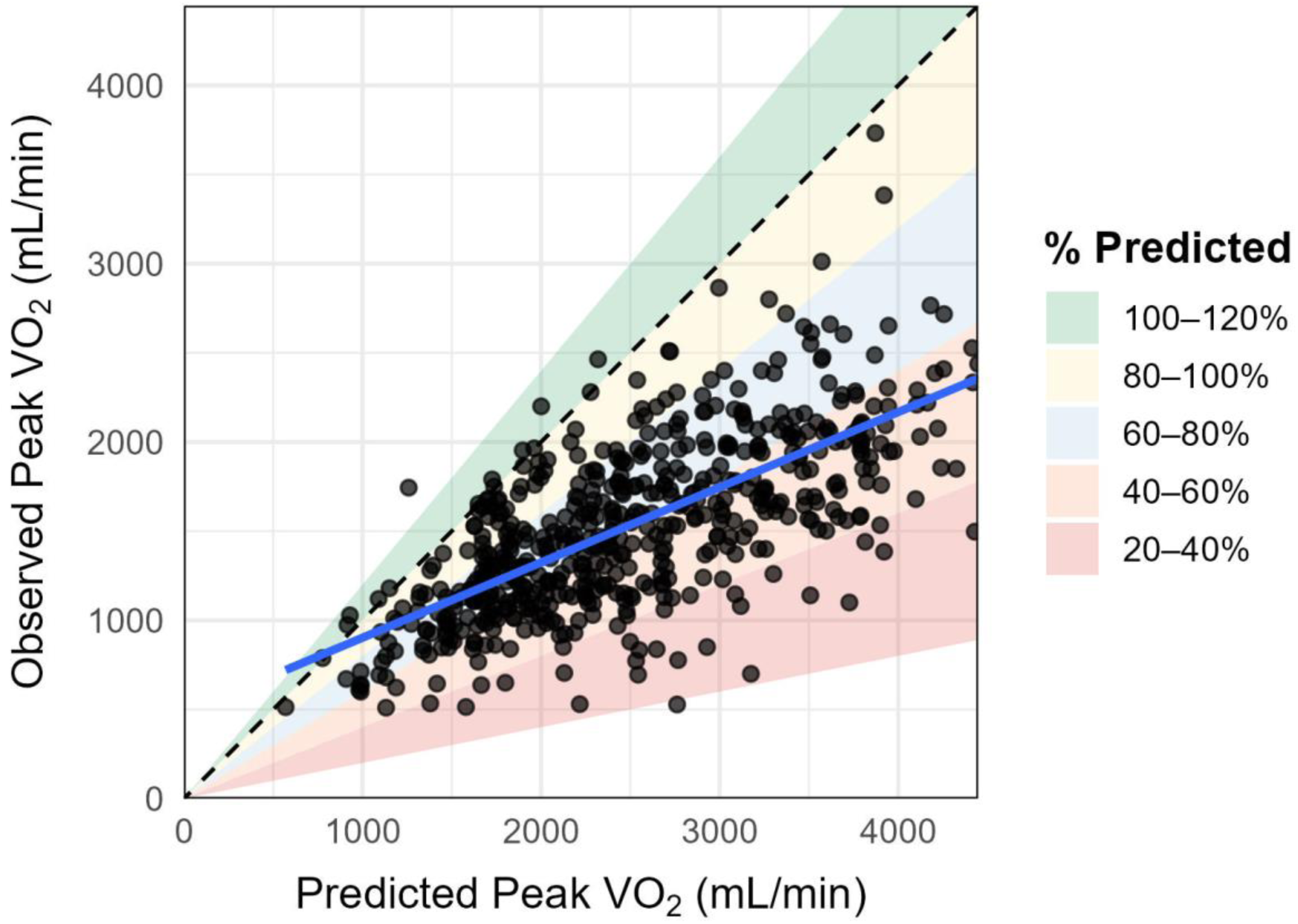
Observed versus predicted peak VO_2_. A **s**catterplot comparing observed peak VO_2_ (mL/min) to predicted peak VO_2_ with the dashed line representing the identity line. Shaded regions denote categories of percent predicted peak VO_2_ (20–40%, 40–60%, 60–80%, 80–100%, and 100–120%). A regression line is fitted (blue) to patients in the cohort. VO_2_ = oxygen consumption.

**Table 2.**
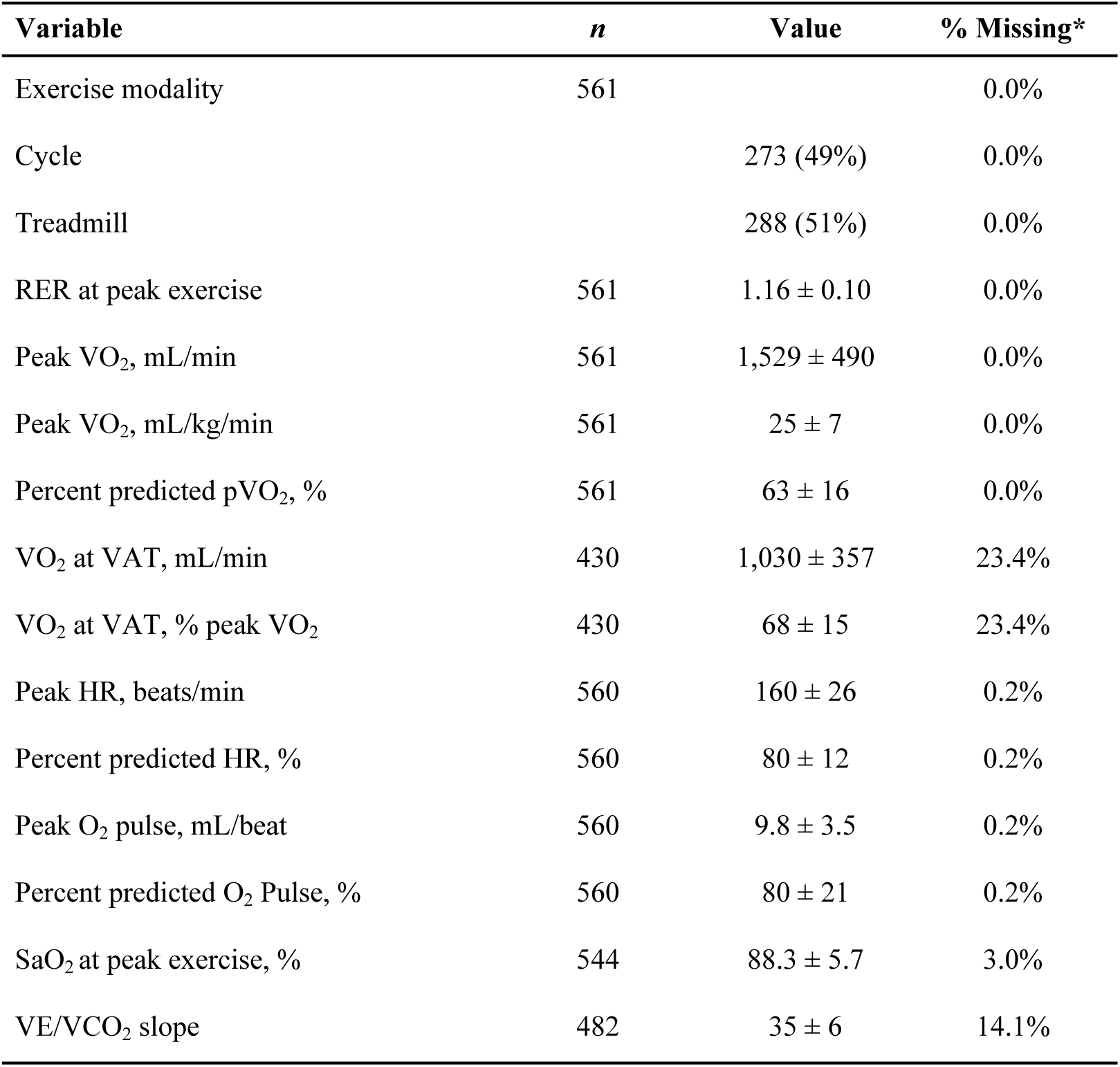
Exercise Data. Data displayed as n (%); mean ± SD. RER = respiratory exchange ratio; VO_2_ = oxygen consumption; pVO_2_ = peak oxygen consumption; VAT = ventilatory anaerobic threshold; HR = heart rate; SaO_2_ = oxygen saturation; VE/VCO_2_ = ventilatory efficiency. *Missing for this table was determined after inclusion/exclusion criteria were applied.

### Non-Parametric and Univariable Modeling

There was a significant difference with respect to sex with females achieving a higher %pVO_2_ than males (p < 0.001). The ECC group had a higher %pVO_2_ than LT (p = 0.02). While the %pVO_2_ for ECC was also higher than APC, it was not significant following Bonferroni adjustment. With respect to ventricular morphology and %pVO_2_, there was no significant difference among the groups (p = 0.42). Percent predicted pVO_2_ did not differ significantly across conduit size categories (p = 0.09). (**Supplemental Material**).

In univariable linear regression models with the outcome variable of %pVO_2_, sex had the highest association with %pVO_2_ with females having a 10.5% higher value than males (p < 0.001, R^2^ = 0.08). For ECC, conduit size indexed to BSA (β = 0.87% per mm/m^2^, p = 0.008, R^2^ = 0.02) was the only conduit size measure that was associated with a higher %pVO_2_. Conduit diameter (β = - 0.6% per mm, p = 0.229, R^2^ = 0.001), conduit area (β = -0.02% per mm^2^, p = 0.165, R^2^ = 0.001), and conduit area indexed to BSA (β = 0.03% per mm^2^, p = 0.086, R^2^ = 0.01) were not associated with exercise capacity measured in %pVO_2_. Comparing across conduit categories, only 18 mm was statistically significant when compared to the 20 mm reference (β = 4.79%, p = 0.015, R^2^ = 0.01).

Patients with balanced or indeterminate ventricular morphology demonstrated slightly higher %pVO_2_ compared with those with right ventricular morphology (β = 8.12%, p = 0.026, R^2^ = 0.01), whereas left ventricular morphology was not associated with higher %pVO_2_. Several demographic factors showed significant relationships with exercise performance: BSA (β = - 8.49% per m^2^, p < 0.001, R^2^ = 0.03), height (β = -0.42% per cm, p < 0.001, R^2^ = 0.08), weight (β = -0.07% per kg, p = 0.039, R^2^ = 0.01), and older age (β = -0.42% per year, p < 0.001, R^2^ = 0.05). Exercise modality (cycle vs treadmill) and initial anatomical diagnoses were not significantly associated with %pVO_2_. In this unadjusted model, the Fontan surgical eras of 1989-1996 and 1997-2005 (when compared with 2006-2013) were associated with worse exercise capacity (β = -5.06, p = 0.039 and -6.65, p < 0.001, respectively; R^2^ = 0.03). These findings are summarized in **Table 3**.

**Table 3.** Univariable Linear Regression Analysis Predicting Percent Predicted Peak VO_2_. For categorical variables, reference groups were chosen as those with the largest sample size. AVSD = atrioventricular septal defect; DORV = double outlet right ventricle; DILV = double inlet left ventricle; HLHS = hypoplastic left heart syndrome; PAIVS = pulmonary atresia with intact ventricular septum; SaO_2_ = oxygen saturation, VO_2_ = oxygen consumption.

| Variable | $\beta$ (95% CI) | p-value | R<sup>2</sup> |
| --- | --- | --- | --- |
| Conduit diameter, mm | -0.6 (-1.57, 0.38) | 0.229 | 0.00 |
| Indexed conduit diameter, mm/m<sup>2</sup> | 0.87 (0.22, 1.51) | **0.008** | 0.02 |
| Conduit area, mm<sup>2</sup> | -0.02 (-0.06, 0.01) | 0.165 | 0.00 |
| Indexed conduit area, mm<sup>2</sup>/m<sup>2</sup> | 0.03 (-0.01, 0.07) | 0.086 | 0.01 |
| Body surface area, m<sup>2</sup> | -8.49 (-12.72, -4.26) | **<0.001** | 0.03 |
| Height, cm | -0.42 (-0.43, -0.24) | **<0.001** | 0.08 |
| Weight, kg | -0.07 (-0.13, -0.01) | **0.039** | 0.01 |
| Age, years | -0.42 (-0.57, -0.27) | **<0.001** | 0.05 |
| Female vs male | 10.5 (7.97, 13.06) | **<0.001** | 0.08 |
| Cycle vs treadmill | -1.65 (-4.33, 1.04) | 0.228 | 0.00 |
| SaO<sub>2</sub> at peak exercise, % | 0.19 (-0.05, 0.43) | 0.125 | 0.00 |
| **Systemic ventricle (ref Right)** |  |  | 0.01 |
| Left | -0.24 (-2.97, 2.49) | 0.861 |  |
| Balanced/indeterminant | 8.12 (0.96, 15.27) | **0.026** |  |
| **Fontan type (ref Extracardiac)** |  |  | 0.01 |
| Lateral tunnel | -4.10 (-7.09, -1.11) | **0.007** |  |
| Atrio-pulmonary connection | -8.06 (-18.70, 2.57) | 0.137 |  |
| Other/unknown | -6.32 (-13.24, 0.60) | 0.073 |  |
| **Conduit diameter (ref = 20 mm)** |  |  | 0.01 |
| ≤14 mm | -2.48 (-15.91, 10.94) | 0.716 |  |
| 16 mm | -0.04 (-6.41, 6.34) | 0.991 |  |
| 18 mm | 4.79 (0.94, 8.63) | 0.015 |  |
| ≥22 mm | -0.87 (-7.73, 5.99) | 0.804 |  |
| **Fontan surgical era (ref 2006-2013)** |  |  | 0.03 |
| 1980-1988 | -3.84 (-17.73, 10.04) | 0.587 |  |
| 1989-1996 | -5.06 (-9.87, -0.25) | **0.039** |  |
| 1997-2005 | -6.65 (-9.98, -3.32) | **<0.001** |  |
| 2014-2022 | 1.80 (-2.09, 5.70) | 0.364 |  |
| **Diagnosis (ref HLHS)** |  |  | 0.00 |
| Balanced AVSD/DORV | -4.78 (-16.26, 6.70) | 0.414 |  |
| DILV | -0.08 (-4.26, 4.09) | 0.969 |  |
| Ebstein | 5.35 (-6.12, 16.83) | 0.360 |  |
| Mitral atresia | -1.37 (-8.67, 5.94) | 0.714 |  |
| Other | 3.46 (-1.50, 8.43) | 0.171 |  |
| PAIVS | -0.25 (-7.01, 6.51) | 0.942 |  |
| Tricuspid atresia | -0.76 (-4.66, 3.15) | 0.704 |  |
| Unbalanced AVSD | -2.17 (-6.63, 2.29) | 0.340 |  |

### Multivariable Linear Regression Modeling

**Table 4** highlights the multivariable model including the surgical, anatomical, and demographic data from 514 patients across the entire cohort to predict %pVO_2_. Female sex remained strongly associated with higher exercise capacity (β = 11.94%, p < 0.001), while older age was associated with lower values (β = -0.80% per year, p < 0.001). Exercise performed on a cycle ergometer was associated with a significantly lower %pVO_2_ compared with treadmill testing (β = -4.59%, p < 0.001). Neither systemic ventricular morphology, anatomical subgroup, nor Fontan subtype showed significant associations after covariate adjustment. The earlier Fontan surgical era of 1989-1996 versus 2006-2013 was associated with higher %pVO_2_ (β = 7.21%, p = 0.018), whereas other eras showed no significant differences. The overall model demonstrated fair explanatory power for the outcome in the setting of this complex clinical phenotype (R^2^ = 0.25, adj. R^2^ = 0.22).

**Table 4.**
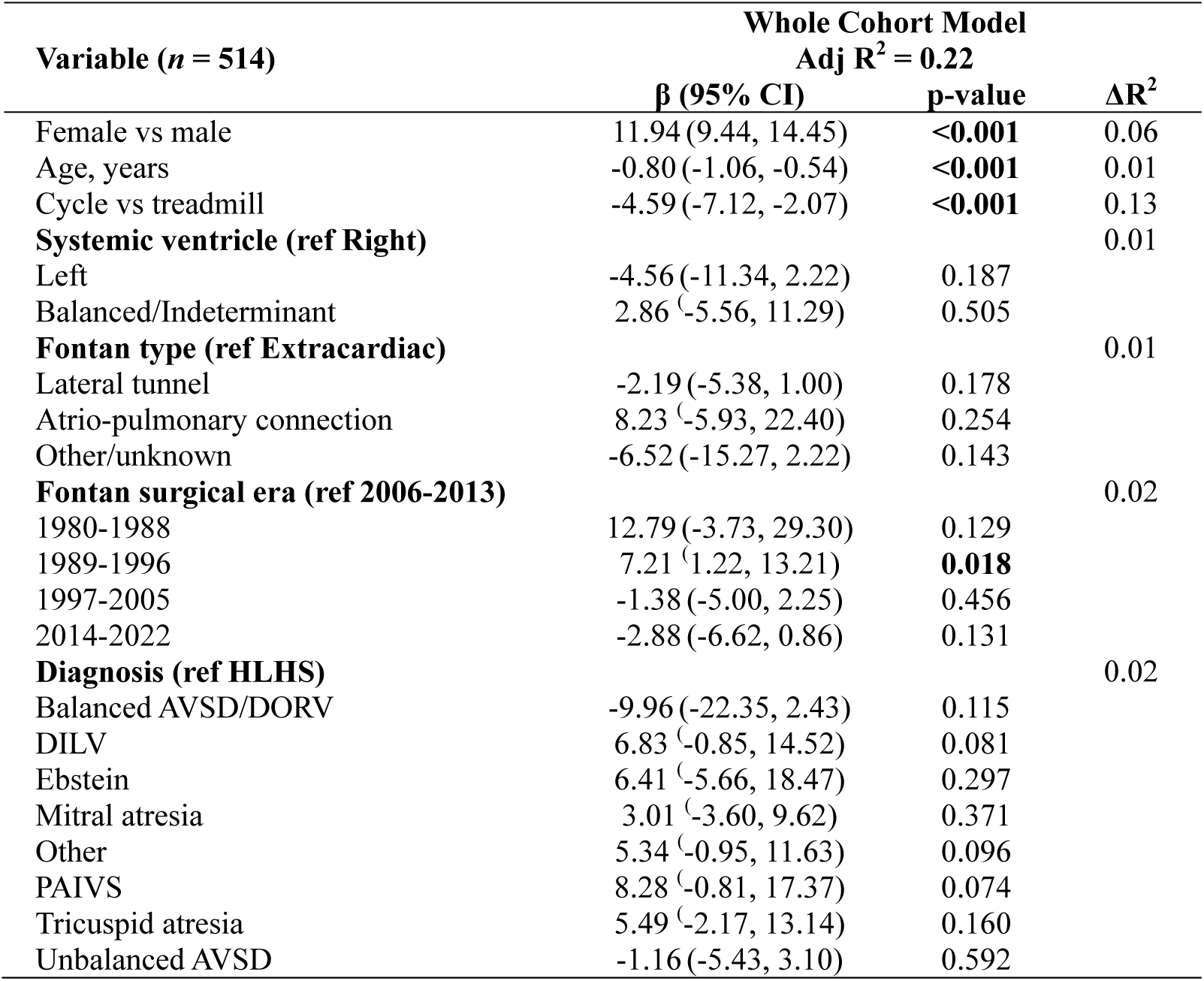
Whole cohort Multivariable Regression Analysis Predicting Percent Predicted Peak VO_2_. This model includes all participants with complete data (n = 514) and evaluates demographic, anatomical, and surgical factors associated with percent predicted peak VO_2_. Reference groups for categorical variables were selected based on the largest sample size. AVSD = atrioventricular septal defect; DORV = double outlet right ventricle; DILV = double inlet left ventricle; HLHS = hypoplastic left heart syndrome; PAIVS = pulmonary atresia with intact ventricular septum; VO_2_ = oxygen consumption; Adj R^2^ = adjusted R^2^. ΔR^2^ = semi-partial R^2^, or each variable’s overall contribution to the model.

Four additional models were constructed to evaluate the independent association between ECC characteristics and %pVO_2_, adjusting for age, sex, exercise modality, ventricular morphology, Fontan surgical era, and diagnosis. There were 330 ECC patients who had data available for all model variables. Across all four models, each of which explored a different conduit size metric, conduit size was not a statistically significant independent predictor of %pVO_2_. Model performance was consistent across specifications, with an adjusted R^2^ of 0.25 for each model. These findings are summarized in **Table 5** with complete regression data in the **Supplemental Material**.

**Table 5.** Multivariable Regression Analysis Among ECC Evaluating the Association Between Conduit Size and Percent Predicted Peak VO_2_. Four separate models were constructed, each incorporating a different conduit measurement (conduit diameter, indexed conduit diameter, conduit area, or indexed conduit area). *These models are adjusted for sex, age, exercise modality, systemic ventricle, Fontan era, and diagnosis. A complete representation of the model variables can be seen in the **Supplemental Material**. VO_2_ = oxygen consumption.

| Conduit Measurement* (*n* = 330) | $\beta$ (95% CI) | p-value |
| --- | --- | --- |
| Diameter (mm) | 0.54 (-0.48, 1.57) | 0.229 |
| Indexed diameter (mm/m<sup>2</sup>) | 0.27 (-0.42, 0.95) | 0.443 |
| Area (mm<sup>2</sup>) | 0.02 (-0.01, 0.06) | 0.226 |
| Indexed area (mm<sup>2</sup>/m<sup>2</sup>) | 0.02 (-0.02, 0.06) | 0.321 |

### Nonlinear Regression for Extracardiac Conduit Size

Nonlinear associations between conduit size and %pVO_2_ were evaluated using univariable restricted cubic spline models. Indexed conduit diameter (mm/m^2^) demonstrated the strongest overall association with %pVO_2_ (p = 0.034), but with only small explanatory power (adj R^2^ = 0.016). The nonlinear term of this relationship was not statistically significant (p = 0.429), indicating that this relationship was predominantly linear across the observed range.

Conduit diameter (mm) and conduit area (mm^2^) were also significantly associated with %pVO_2_ (diameter: p = 0.042, adj R^2^ = 0.015; area: p = 0.040, adj R^2^ = 0.015). For conduit diameter (mm), the spline suggested a peak in %pVO_2_ at 18 mm, with diminishing returns at larger diameters (20–22 mm), consistent with a nonlinear relationship. In contrast, indexed conduit area (mm^2^/m^2^) did not have a significant relationship (p = 0.338). These plots are illustrated in **Figure 3**.

**Figure 3.**
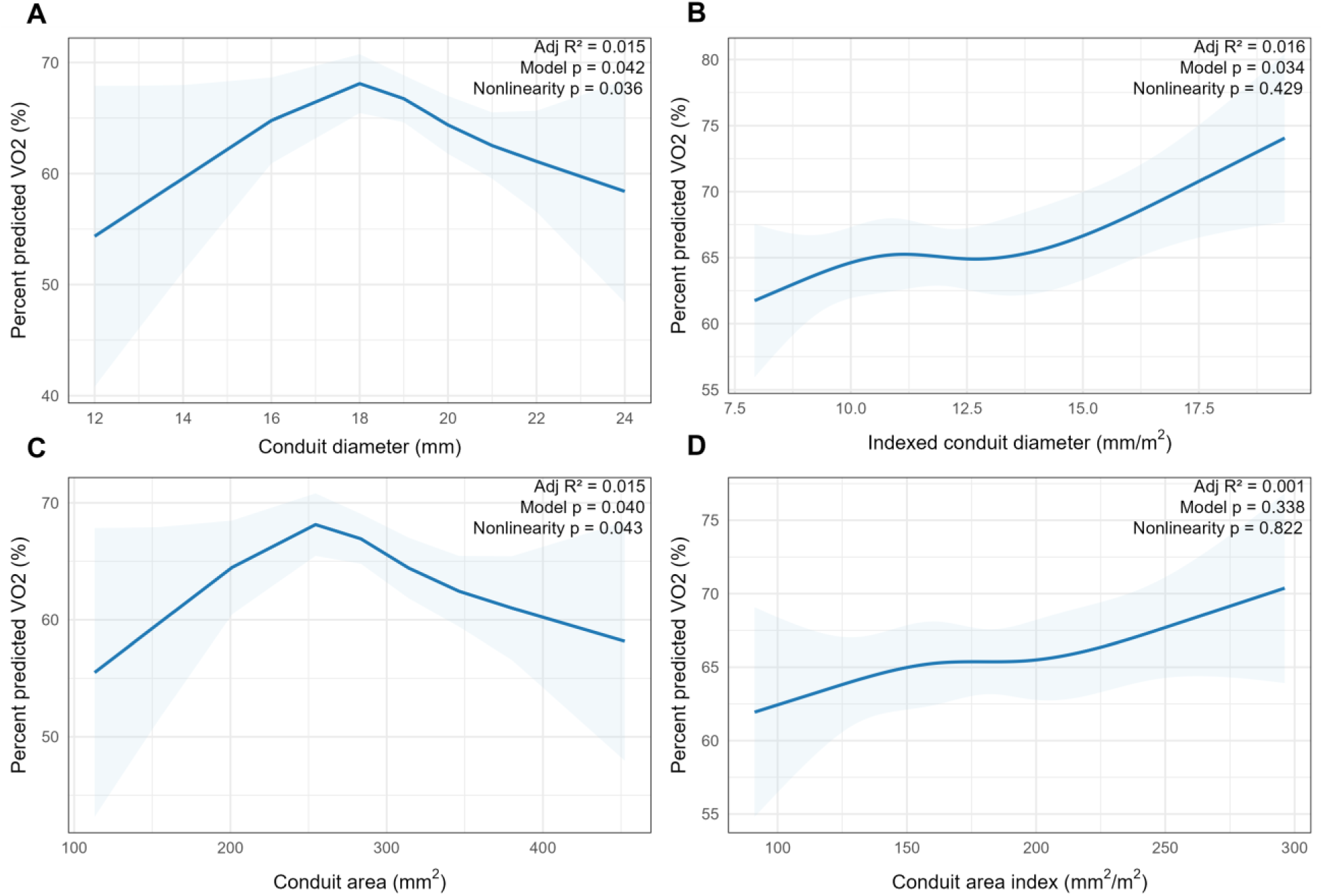
Univariable Nonlinear Spline Plots for Conduit Size Measures and Percent Predicted Peak VO_2_. Panel A shows the relationship between absolute conduit diameter and percent predicted peak VO_2_. Panel B displays the relationship for conduit diameter indexed to body surface area. Panel C depicts absolute conduit area, while Panel D shows conduit area indexed to body surface area. Shaded regions represent model-derived confidence intervals. VO_2_ = oxygen consumption.

### Sensitivity Analyses

Sensitivity analysis was performed using the Wasserman/Hansen equation.^29^ Compared to the FRIEND equation, Wasserman/Hansen estimated a lower pVO_2_ (and thus higher %pVO_2_). When comparing models using each estimate for adults, the FRIEND equation had an improved fit (via R^2^) across all models and a lower standard deviation. Thus, FRIEND was utilized as the primary equation for adults in the analysis.

Analysis was also performed on the ECC models using only 18 vs 20 mm conduits (*n* = 273) to determine if there was an effect of these particular sizes on %pVO_2_. The results of this modeling showed no difference in %pVO_2_ between 18 vs 20 mm (β = 0.40%, p = 0.829) after controlling for covariates. Sensitivity analysis data are available in the **Supplemental Material**.

## DISCUSSION

This study leveraged a large multicenter dataset using the FON registry, now within SV-ONE, and represents one of the largest evaluations of CPET data in patients with a Fontan circulation. Conduit size at the time of Fontan creation was not associated with higher exercise capacity later in life when controlling for other important factors of exercise performance, such as body size, sex, and modality. Although %pVO_2_ equations account for age and sex, both variables remained significant predictors of %pVO_2_ in all models, suggesting that population reference equations differ from patterns observed in patients with a Fontan circulation. Sex and exercise modality had the largest effect on predicting %pVO_2_ with females demonstrating on average a 12% higher %pVO_2_ than males, consistent with prior reports by others in patients following Fontan surgery.^10,30,31^ Fontan subtype was not independently associated with %pVO_2_, nor was ventricular morphology or underlying diagnosis. As an overall cohort, there was a reduced exercise capacity compared with population norms, consistent with prior reports.^3,15,32,33^

### Implanted Conduit Size

Conduit size and area were not associated with a change in %pVO_2_. While indexed conduit diameter (mm/m^2^) reached statistical significance, this effect was small and likely reflects confounding by body size, as BSA, height, sex, and age were stronger univariable predictors of %pVO_2_. This was confirmed in multivariable analysis. Overall, these findings suggest that conduit size itself is not a primary determinant of later exercise performance.

We also conducted a nonlinear assessment to identify a possibly optimal conduit size. Modeling suggested that the 18 mm conduit (260 mm^2^) was optimal. Similar nonlinear patterns between indexed conduit size and exercise capacity have been reported previously.^34^ When the conduit size was indexed to body size (panels B and D of **Figure 3**), the statistical significance in the model was no longer observed.

Taken together, these findings suggest that exercise limitation in the Fontan circulation may be driven predominantly by factors other than conduit dimensions, such as pulmonary vascular resistance, ventricular systolic reserve during exercise, impaired preload augmentation, and elevated single-ventricle filling pressures.^35,36^ In addition, initial conduit size has been shown to influence other clinically relevant outcomes not assessed in this study, such as hepatic or renal congestion, and does not capture abnormalities in postoperative pulmonary blood flow patterns, both of which warrant further investigation.^37,38^ There is also concern that conduit size may decrease with time, highlighting the distinction between implanted conduit size and contemporaneous luminal dimensions assessed at the time of exercise testing.^18^

### Ventricular Morphology

A longstanding clinical question is whether a systemic right ventricle, which is not designed to sustain systemic afterload, is associated with worse exercise performance in patients with a Fontan circulation. Prior studies have reported higher %pVO2 in those with a systemic left ventricle.^11,39^ In contrast, we did not observe a significant difference in exercise capacity between left and right ventricular morphology. This discrepancy may relate to differences in cohort age, as adverse effects of a systemic right ventricle may become more apparent later in life, a pattern well described in adults with transposition of the great arteries repaired with atrial switch procedures.^40,41^

### Fontan Subtype and Surgical Era

We examined whether Fontan subtype or surgical era was independently associated with exercise performance. Although unadjusted differences in %pVO_2_ have been reported between extracardiac conduit and lateral tunnel configurations, these associations typically attenuate after adjustment for physiologic and demographic factors. Consistent with prior data, Fontan subtype was not independently associated with %pVO_2_ in our fully adjusted whole-cohort model.^42^

Surgical era showed an association only for the 1989–1996 group compared with the 2006–2013 reference group, without a clear mechanistic explanation and with potential confounding from survival bias and age-related cohort effects. Overall, these findings support that exercise capacity in patients with a Fontan circulation is driven more by demographic and physiologic factors than by Fontan configuration or operative era.

### Clinical Context

A strength of this study is the inclusion of a broad age range, enabling characterization of age-related differences in exercise capacity among patients with a Fontan circulation. Together with recent multicenter registry data, our findings suggest that long-term outcomes are more strongly influenced by age-related trajectories, exercise participation, muscle strength, ventricular and valvular function, and pulmonary vascular physiology than by surgical factors, underscoring the need for age-specific benchmarks when interpreting CPET data.^11,14,20,35,39,43,44^ Future studies should move beyond conduit diameter to incorporate Fontan pathway geometry, pulmonary artery abnormalities, hemodynamics, biomarkers, and serial exercise testing to better define functional phenotypes and refine risk stratification across the lifespan. The basis for observed sex differences also warrants further investigation.

### Limitations

As with any registry-based study, this cohort is subject to potential selection bias at both the center and patient levels, though likely less than in single-center studies. Although we adjusted for major confounders, unmeasured factors such as center-specific practices, surgical technique, and postoperative management were not captured. Data quality depended on clinical documentation and site-level entry, introducing possible misclassification or missingness, particularly for conduit characteristics and CPET variables. Importantly, %pVO_2_ and other CPET metrics may not capture all potential benefits of larger conduit size; for example, a larger conduit could be associated with lower venous pressures and reduced cumulative end-organ injury independent of exercise capacity.

Conduit diameter and area were based on nominal implanted size and assumed circular geometry, reflecting surgical selection but not necessarily true luminal dimensions or geometry at CPET. Interval changes such as kinking, distortion, stenosis, transcatheter interventions, or branch pulmonary artery narrowing were not captured and may influence pulmonary blood flow and exercise performance. Residual confounding cannot be excluded, and longitudinal studies are needed to better define individual functional trajectories. Indexing to body surface area at the time of CPET rather than at Fontan completion may also affect interpretation. Although VO_2_ reference equations provide a practical method for indexing to multiple physiologic parameters, they have recognized limitations, including age-related assumptions, methodological variability across cohorts, and potential selection bias. Finally, survival bias is possible, particularly among older individuals, as those who underwent CPET may represent a healthier subset.

## CONCLUSION

In this large multicenter cohort, early anatomical and surgical features were not strong determinants of later exercise performance. Although exercise capacity was broadly reduced compared with population norms, this likely reflects the complex, progressively maladaptive physiology of the Fontan circulation rather than static surgical characteristics alone. Exercise performance represents only one dimension of Fontan health and does not fully capture other important outcomes such as arrhythmia burden, end-organ function, symptom burden, or quality of life. Overall, long-term status in patients with a Fontan circulation appears to be driven more by age-related changes and downstream physiological processes than by anatomical factors alone, supporting a longitudinal, multidimensional approach to assessment and risk stratification across the lifespan.

## Data Availability

Procedure for Sharing Data (For principal investigators not from a FON Member Center) 1.External researcher submits proposed research question(s) and analytic methods plan to Research Workgroup for preliminary approval. The question and plan are reviewed based on scientific rigor and if the research question can be answered using the FON database. 2. If the proposal receives preliminary approval: a. The FON Research Workgroup Representative sends the Data Request Form to the researcher to sign and send back. b. Researcher submits full proposal to their own IRB for approval as well as to the FON Research Workgroup Representative who will review it to ensure it is keeping to the community norms. 3. If proposal is approved by the researcher?s IRB and FON Research Workgroup Representative: a. Researcher submits IRB approval letter to FON Research Workgroup b. FON Research Workgroup Representative notifies Data Workgroup of IRB approval and any other needed approvals.

https://www.svone.org/

## ACKNOWLEDGMENT

The ideas expressed in this manuscript do not represent formal positions of the Single Ventricle Outcomes Network (SV-ONE). We would like to thank the registrants and the sites for collecting data.

The authors used generative artificial intelligence (ChatGPT 5.2; OpenAI) as an aid in writing code for data analysis and in editing and proofreading the manuscript. All output from this tool has been reviewed, edited, and modified, as needed by the authors and the authors assume full responsibility for the accuracy and integrity of the scientific content presented in this publication.

## SUPPLEMENTAL MATERIAL

Tables S1 & S2

Figures S1-S8

SV-ONE Investigators Supplemental Appendix

## ABBREVIATIONS

APC: Atrio-pulmonary connection
BSA: Body surface area
CPET: Cardiopulmonary exercise testing
ECC: Extracardiac conduit
FRIEND: Fitness Registry and the Importance of Exercise: A National Database
FON: Fontan Outcomes Network
HR: Heart rate
LT: Lateral tunnel
NPC-QIC: National Pediatric Cardiology Quality Improvement Collaborative
pVO_2_: Peak oxygen consumption
%pVO_2_: Percent predicted peak oxygen consumption
RER: Respiratory exchange ratio
SV-ONE: Single Ventricle Outcomes Network
VAT: Ventilatory anaerobic threshold
VE/VCO_2_: Ratio of minute ventilation to carbon dioxide production

## REFERENCES

1. Downing KF, Nembhard WN, Rose CE, Andrews JG, Goudie A, Klewer SE, Oster ME, Farr SL. Survival From Birth Until Young Adulthood Among Individuals With Congenital Heart Defects: CH STRONG. Circulation. 2023;148:575–588. doi: 10.1161/circulationaha.123.064400

2. Budts W, Ravekes WJ, Danford DA, Kutty S. Diastolic Heart Failure in Patients With the Fontan Circulation: A Review. JAMA Cardiol. 2020;5:590–597. doi: 10.1001/jamacardio.2019.5459

3. Kempny A, Dimopoulos K, Uebing A, Moceri P, Swan L, Gatzoulis MA, Diller GP. Reference values for exercise limitations among adults with congenital heart disease. Relation to activities of daily life--single centre experience and review of published data. Eur Heart J. 2012;33:1386–1396. doi: 10.1093/eurheartj/ehr461

4. Egbe AC, Driscoll DJ, Khan AR, Said SS, Akintoye E, Berganza FM, Connolly HM. Cardiopulmonary exercise test in adults with prior Fontan operation: the prognostic value of serial testing. International journal of cardiology. 2017;235:6–10.

5. Lala A, Shah KB, Lanfear DE, Thibodeau JT, Palardy M, Ambardekar AV, McNamara DM, Taddei-Peters WC, Baldwin JT, Jeffries N, et al. Predictive Value of Cardiopulmonary Exercise Testing Parameters in Ambulatory Advanced Heart Failure. JACC Heart Fail. 2021;9:226–236. doi: 10.1016/j.jchf.2020.11.008

6. Lewis GD, Zlotoff DA. Cardiopulmonary Exercise Testing-Based Risk Stratification in the Modern Era of Advanced Heart Failure Management. JACC Heart Fail. 2021;9:237–240. doi: 10.1016/j.jchf.2021.01.003

7. Agarwal A, Cunnington C, Sabanayagam A, Zier L, McCulloch CE, Harris IS, Foster E, Atkinson D, Bryan A, Jenkins P, et al. Cardiopulmonary exercise testing in the evaluation of liver disease in adults who have had the Fontan operation. Arch Cardiovasc Dis. 2018;111:276–284. doi: 10.1016/j.acvd.2017.09.001

8. Inuzuka R, Diller GP, Borgia F, Benson L, Tay EL, Alonso-Gonzalez R, Silva M, Charalambides M, Swan L, Dimopoulos K, et al. Comprehensive use of cardiopulmonary exercise testing identifies adults with congenital heart disease at increased mortality risk in the medium term. Circulation. 2012;125:250–259. doi: 10.1161/CIRCULATIONAHA.111.058719

9. Cordina R, du Plessis K, Tran D, d’Udekem Y. Super-Fontan: Is it possible? J Thorac Cardiovasc Surg. 2018;155:1192–1194. doi: 10.1016/j.jtcvs.2017.10.047

10. Tran DL, Celermajer DS, Ayer J, Grigg L, Clendenning C, Hornung T, Justo R, Davis GM, d’Udekem Y, Cordina R. The “Super-Fontan” Phenotype: Characterizing Factors Associated With High Physical Performance. Front Cardiovasc Med. 2021;8:764273. doi: 10.3389/fcvm.2021.764273

11. Alsaied T, Li R, Christopher AB, Fogel MA, Slesnick TC, Krishnamurthy R, Muthurangu V, Dorfman AL, Lam CZ, Weigand JD, et al. High-Performing Fontan Patients: A Fontan Outcome Registry by Cardiac Magnetic Resonance Imaging Study. JACC Adv. 2024;3:101254. doi: 10.1016/j.jacadv.2024.101254

12. Dalén M, Odermarsky M, Liuba P, Johansson Ramgren J, Synnergren M, Sunnegårdh J. Long-Term Survival After Single-Ventricle Palliation: A Swedish Nationwide Cohort Study. J Am Heart Assoc. 2024;13:e031722. doi: 10.1161/jaha.123.031722

13. Dhauna J, Aboulhosn J, Lluri G. Cardiopulmonary Exercise Test Outcomes in Fontan Patients With Right Versus Left Single Ventricle Morphology. World J Pediatr Congenit Heart Surg. 2022;13:366–370. doi: 10.1177/21501351221087695

14. Seese L, Schiff M, Olivieri L, Da Fonseca Da Silva L, Da Silva JP, Christopher A, Harris TH, Morell V, Castro Medina M, Rathod RH, et al. Differences in Exercise Performance in Fontan Patients with Extracardiac Conduit and Lateral Tunnel: A FORCE Fontan Registry Study. J Clin Med. 2025;14. doi: 10.3390/jcm14124067

15. Paridon SM, Mitchell PD, Colan SD, Williams RV, Blaufox A, Li JS, Margossian R, Mital S, Russell J, Rhodes J. A cross-sectional study of exercise performance during the first 2 decades of life after the Fontan operation. Journal of the American College of Cardiology. 2008;52:99–107. doi: 10.1016/j.jacc.2008.02.081

16. Mays WA, Border WL, Knecht SK, Gerdes YM, Pfriem H, Claytor RP, Knilans TK, Hirsch R, Mone SM, Beekman RH, 3rd. Exercise capacity improves after transcatheter closure of the Fontan fenestration in children. Congenit Heart Dis. 2008;3:254–261. doi: 10.1111/j.1747-0803.2008.00199.x

17. Ortega-Aviles LM, Santos-Patarroyo SD, Opp DN, Cetta F, Egbe AC, Miranda WR, Allison TG. Cardiopulmonary exercise test variations in patients with a Fontan circulation: impact of fenestration patency and associations with clinical outcomes. Open Heart. 2025;12. doi: 10.1136/openhrt-2025-003639

18. Patel ND, Friedman C, Herrington C, Wood JC, Cheng AL. Progression in Fontan conduit stenosis and hemodynamic impact during childhood and adolescence. J Thorac Cardiovasc Surg. 2021;162:372–380.e372. doi: 10.1016/j.jtcvs.2020.09.140

19. Bove EL, de Leval MR, Migliavacca F, Guadagni G, Dubini G. Computational fluid dynamics in the evaluation of hemodynamic performance of cavopulmonary connections after the Norwood procedure for hypoplastic left heart syndrome. J Thorac Cardiovasc Surg. 2003;126:1040–1047. doi: 10.1016/s0022-5223(03)00698-6

20. Meschin JB, Mai A, Wang-Giuffre E. Predictors of Superior Exercise Performance in Patients Following Fontan Palliation. Pediatric cardiology. 2025. doi: 10.1007/s00246-025-04129-4

21. Stöcker F, Neidenbach R, Fritz C, Oberhoffer RM, Ewert P, Hager A, Nagdyman N. Oxygen Availability in Respiratory Muscles During Exercise in Children Following Fontan Operation. Front Pediatr. 2019;7:96. doi: 10.3389/fped.2019.00096

22. Carey PM, Yeh HW, Krzywda K, Teson KM, Watson JS, Goudar S, Forsha D, White DA. Moderators of peak respiratory exchange ratio during exercise testing in children and adolescents with Fontan physiology. Cardiol Young. 2023;33:2334–2341. doi: 10.1017/s1047951123000227

23. Chase PJ, Kenjale A, Cahalin LP, Arena R, Davis PG, Myers J, Guazzi M, Forman DE, Ashley E, Peberdy MA, et al. Effects of respiratory exchange ratio on the prognostic value of peak oxygen consumption and ventilatory efficiency in patients with systolic heart failure. JACC Heart Fail. 2013;1:427–432. doi: 10.1016/j.jchf.2013.05.008

24. de Souza ESCG, Kaminsky LA, Arena R, Christle JW, Araújo CGS, Lima RM, Ashley EA, Myers J. A reference equation for maximal aerobic power for treadmill and cycle ergometer exercise testing: Analysis from the FRIEND registry. Eur J Prev Cardiol. 2018;25:742–750. doi: 10.1177/2047487318763958

25. Cooper DM, Weiler-Ravell D. Gas exchange response to exercise in children. Am Rev Respir Dis. 1984;129:S47–48. doi: 10.1164/arrd.1984.129.2P2.S47

26. Egbe AC, Ali AE, Miranda WR, Connolly HM, Borlaug BA. Aerobic Capacity of Adults With Fontan Palliation: Disease-Specific Reference Values and Relationship to Outcomes. Circ Heart Fail. 2025;18:e011981. doi: 10.1161/circheartfailure.124.011981

27. Myers J, de Souza ESCG, Arena R, Kaminsky L, Christle JW, Busque V, Ashley E, Moneghetti K. Comparison of the FRIEND and Wasserman-Hansen Equations in Predicting Outcomes in Heart Failure. J Am Heart Assoc. 2021;10:e021246. doi: 10.1161/jaha.121.021246

28. von Elm E, Altman DG, Egger M, Pocock SJ, Gøtzsche PC, Vandenbroucke JP. Strengthening the Reporting of Observational Studies in Epidemiology (STROBE) statement: guidelines for reporting observational studies. Bmj. 2007;335:806–808. doi: 10.1136/bmj.39335.541782.AD

29. Hansen JE, Sue DY, Wasserman K. Predicted values for clinical exercise testing. Am Rev Respir Dis. 1984;129:S49–55. doi: 10.1164/arrd.1984.129.2P2.S49

30. Goldberg DJ, Zak V, McCrindle BW, Ni H, Gongwer R, Rhodes J, Garofano RP, Kaltman JR, Lambert LM, Mahony L, et al. Exercise Capacity and Predictors of Performance After Fontan: Results from the Pediatric Heart Network Fontan 3 Study. Pediatric cardiology. 2021;42:158–168. doi: 10.1007/s00246-020-02465-1

31. Gurvitz M, Krieger EV, Fuller S, Davis LL, Kittleson MM, Aboulhosn JA, Bradley EA, Buber J, Daniels CJ, Dimopoulos K, et al. 2025 ACC/AHA/HRS/ISACHD/SCAI Guideline for the Management of Adults With Congenital Heart Disease: A Report of the American College of Cardiology/American Heart Association Joint Committee on Clinical Practice Guidelines. Journal of the American College of Cardiology. 2025. doi: 10.1016/j.jacc.2025.09.006

32. Cunningham JW, Nathan AS, Rhodes J, Shafer K, Landzberg MJ, Opotowsky AR. Decline in peak oxygen consumption over time predicts death or transplantation in adults with a Fontan circulation. American heart journal. 2017;189:184–192.

33. Goldberg DJ, Avitabile CM, McBride MG, Paridon SM. Exercise capacity in the Fontan circulation. Cardiology in the Young. 2013;23:824–830. doi: 10.1017/s1047951113001649

34. Lee SY, Song MK, Kim GB, Bae EJ, Kim SH, Jang SI, Cho SK, Kawk JG, Kim WH, Lee CH, et al. Relation Between Exercise Capacity and Extracardiac Conduit Size in Patients with Fontan Circulation. Pediatric cardiology. 2019;40:1584–1590. doi: 10.1007/s00246-019-02190-4

35. Egbe AC, Connolly HM, Miranda WR, Ammash NM, Hagler DJ, Veldtman GR, Borlaug BA. Hemodynamics of Fontan Failure: The Role of Pulmonary Vascular Disease. Circ Heart Fail. 2017;10. doi: 10.1161/circheartfailure.117.004515

36. Egbe AC, Miranda WR, Anderson JH, Borlaug BA. Hemodynamic and Clinical Implications of Impaired Pulmonary Vascular Reserve in the Fontan Circulation. Journal of the American College of Cardiology. 2020;76:2755–2763. doi: 10.1016/j.jacc.2020.10.003

37. Marathe SP, Betts KS, Venna A, Daley M, Iyengar AJ, Cordina R, Celermajer D, Andrews D, Robertson T, Liava’a M, et al. Development of a patient-specific Fontan failure risk calculator using machine learning-a step toward personalized medicine. J Thorac Cardiovasc Surg. 2026. doi: 10.1016/j.jtcvs.2025.12.032

38. Ponzoni M, Saleh J, Chaturvedi RR, Dipchand AI, Valverde I, Seed M, Yoo SJ, Coles J, Honjo O, Lam CZ. Extracardiac conduit restriction is associated with increased liver fibrosis in adolescent Fontan patients. J Thorac Cardiovasc Surg. 2026;171:328–337.e325. doi: 10.1016/j.jtcvs.2025.09.044

39. Anderson-Bell DM, Hardison EH, Rashid M, Hammond BH, Hegewald M, Rapp TE, Ploutz M, Williams RV, Ziebell D, Chaiyakunapruk N. Fontan Patients with a Systemic Left Ventricle Have Greater Exercise Capacity than those with a Systemic Right Ventricle: A Systematic Review and Meta-Analysis. Pediatric cardiology. 2025. doi: 10.1007/s00246-025-03932-3

40. Cuypers JA, Eindhoven JA, Slager MA, Opić P, Utens EM, Helbing WA, Witsenburg M, van den Bosch AE, Ouhlous M, van Domburg RT, et al. The natural and unnatural history of the Mustard procedure: long-term outcome up to 40 years. Eur Heart J. 2014;35:1666–1674. doi: 10.1093/eurheartj/ehu102

41. Ghelani SJ, Lu M, Sleeper LA, Prakash A, Castellanos DA, Clair NS, Powell AJ, Rathod RH. Longitudinal changes in ventricular size and function are associated with death and transplantation late after the Fontan operation. J Cardiovasc Magn Reson. 2022;24:56. doi: 10.1186/s12968-022-00884-y

42. Bossers SS, Helbing WA, Duppen N, Kuipers IM, Schokking M, Hazekamp MG, Bogers AJ, Ten Harkel AD, Takken T. Exercise capacity in children after total cavopulmonary connection: lateral tunnel versus extracardiac conduit technique. J Thorac Cardiovasc Surg. 2014;148:1490–1497. doi: 10.1016/j.jtcvs.2013.12.046

43. Wittekind SG, Powell AW, Opotowsky AR, Mays WW, Knecht SK, Rivin G, Chin C. Skeletal Muscle Mass Is Linked to Cardiorespiratory Fitness in Youth. Med Sci Sports Exerc. 2020;52:2574–2580. doi: 10.1249/mss.0000000000002424

44. Lui MC, Palaniappan L, Leonard MB, Long J, Dehoney J, Cooke JP, Olson I, Damase TR, Chen S, Myers J, et al. Exercise Capacity, Endothelial Function, Muscle Mass, and Strength in Pediatric Patients With Fontan Circulation. CJC Pediatric and Congenital Heart Disease. 2025. doi: 10.1016/j.cjcpc.2025.10.010

